# Evidence that higher temperatures are associated with lower incidence of COVID-19 in pandemic state, cumulative cases reported up to March 27, 2020

**DOI:** 10.1101/2020.04.02.20051524

**Authors:** Michael Triplett

## Abstract

Seasonal temperature variation may impact the trajectories of COVID-19 in different global regions. Cumulative data reported by the World Health Organization, for dates up to March 27, 2020^1^, show association between COVID-19 incidence and regions at or above 30° latitude. Historic climate data also show significant reduction of case rates with mean maximum temperature above approximately 22.5 degrees Celsius. Variance at the local level, however, could not be well explained by geography and temperature. These preliminary findings support continued countermeasures and study of SARS-CoV-2/COVID-19 transmission rates by temperature and humidity.

## Background

In recent months, pandemic COVID-19, which is caused by a coronavirus called SARS-CoV-2 that originated in Wuhan, China, has spread rapidly across temperate regions of the northern hemisphere^6^. Concurrent speculation has grown about the trajectory of the virus in various regions. On March 30, 2020 Dr. Marc Lipsitch, Professor of Epidemiology at Harvard University, said, “It’s really clear that warmer weather does not stop the transmission or growth of the virus… There’s no question that coronaviruses are capable of transmitting in hotter, humid climates.”^**10**^ However, several early studies, yet to be peer reviewed at the time of this research, indicate reduced transmission with increased temperature^**4,6,7**^. Some respiratory conditions, like seasonal influenza, are already known to show consistent trends. Study of SARS coronavirus, which broke out in Hong Kong in 2003, has also shown dependence on temperature and humidity^**5,8**^. This study aims to provide data supporting further research of the effects of temperature and humidity on COVID-19/SARS-CoV-2.

## Materials and Methods

Daily COVID-19 situation reports provided by the World Health Organization (WHO) were obtained for dates beginning March 14 and ending March 27^1^. Information included cumulative case data for all reporting nations. Confirmed case counts were recorded for all nations for the dates listed. Population data assigned to reporting nations were taken from UN estimates of “Total population, both sexes combined” (UNdata, United Nations Statistics Division, 17 June 2019) and used to calculate confirmed case rates (confirmed cases / population). Nations showing case rates above 0.1% with populations less than 1 million were treated as outliers and omitted from analysis. Nations without available predictor data (i.e. population) were also omitted from analysis.

Population values were assigned locations for case-mapping by taking the average (middle) of each nation’s most extreme latitudes and longitudes (i.e. the latitudes and longitudes of each nation’s northern-, eastern-, southern- and western-most points). For nations in the western hemisphere above 40 degrees latitude at center, and for nations in the eastern hemisphere above 60 degrees latitude at center, populations were assumed to be concentrated near the southernmost border. Fig. 1 shows how populations were represented from -65° to 65° degrees latitude

**Figure 1:**
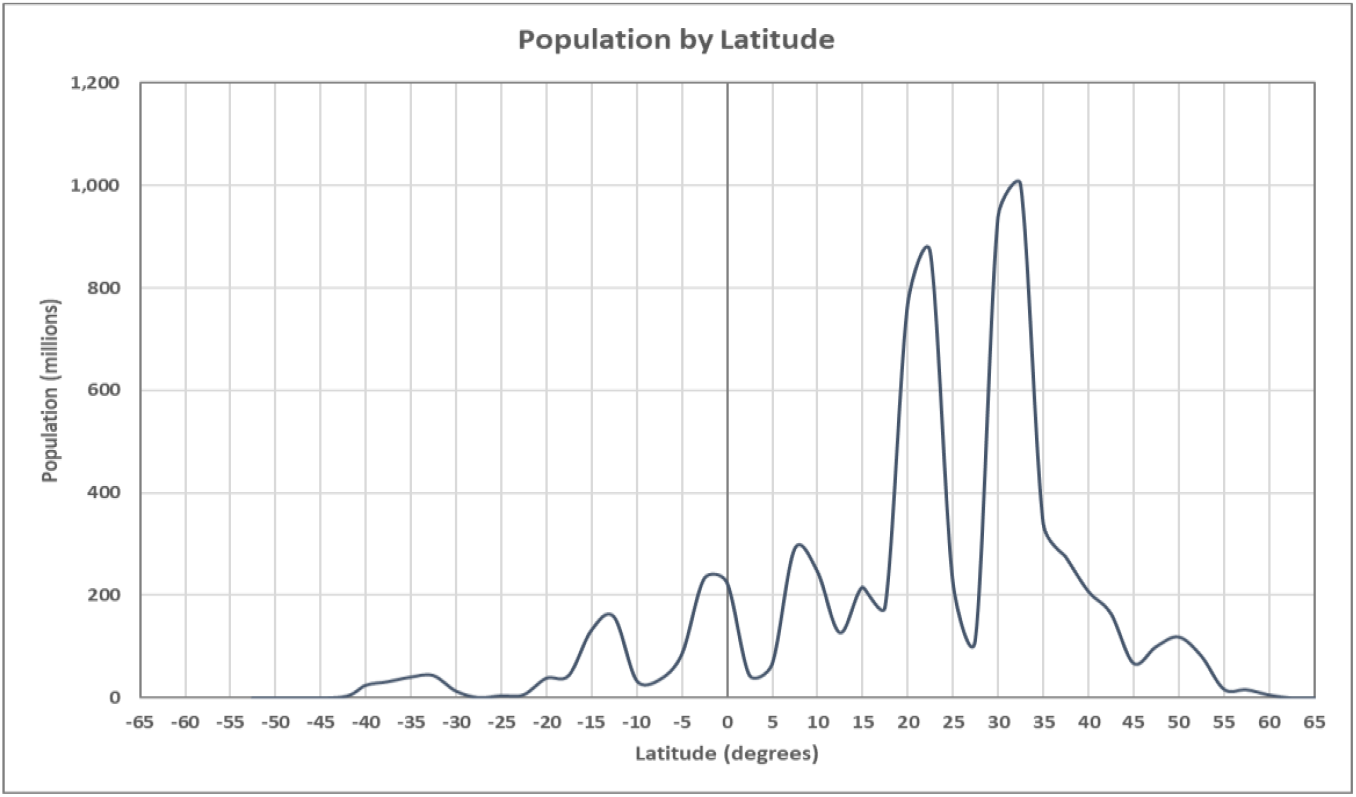
Global population as modeled by assigned latitude

Global gridded daily maximum surface temperature data, with 0.5° spatial resolution, for the dates from February 29 to March 14, 2020, were accessed from the US National Oceanic and Atmospheric Administration (NOAA), Earth system research Laboratory (ESRL), Physical Sciences Division website (ftp://ftp.cdc.noaa.gov/Datasets/cpc_global_temp/)^3^. Dates two weeks prior to respective case dates were chosen to account for 14 days between transmission and case confirmation. As shown in fig. 2, recorded mean maximum temperature values were averaged across each reported latitude for each day and assigned to reporting nations based on modeled population latitudes.

**Figure 2:**
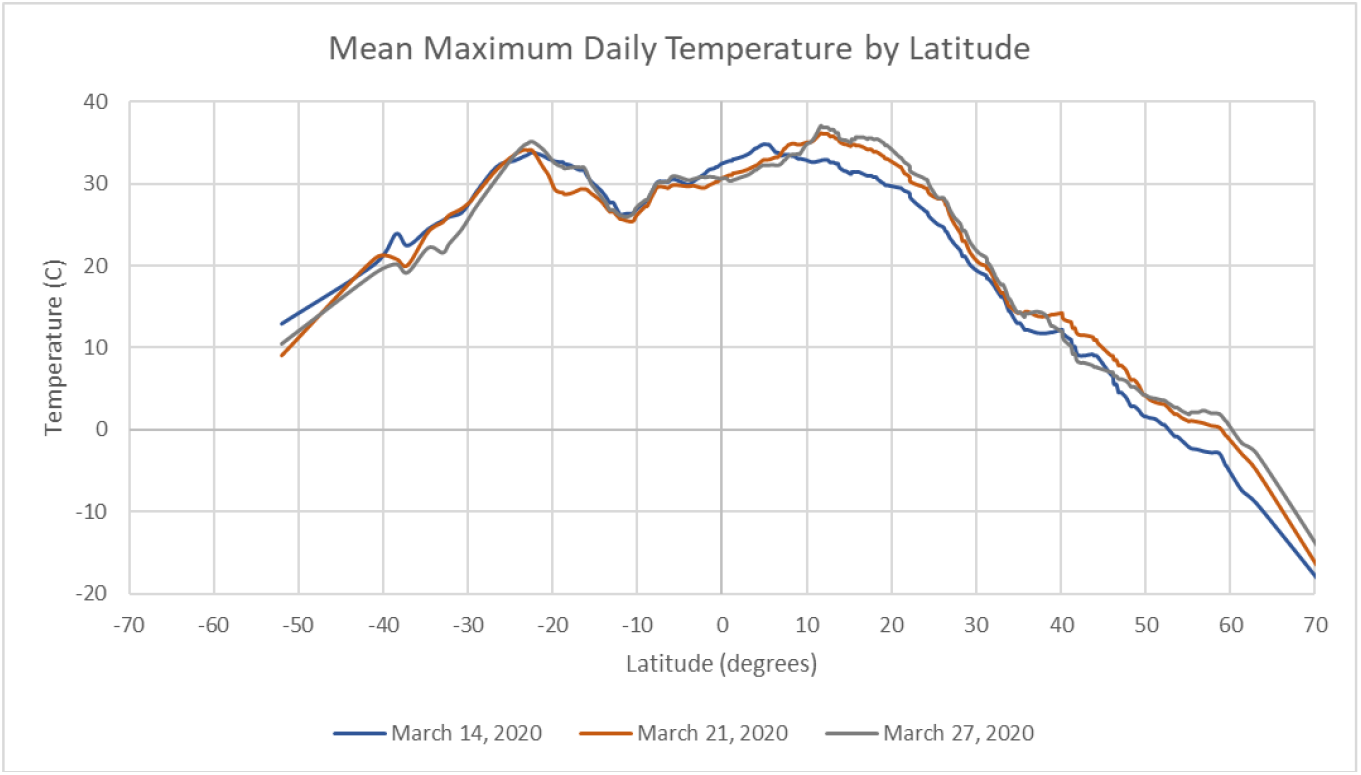
Mean maximum surface temperature by latitude for March 14, March 21 and March 27, 2020

For data reported March 27, 2020, multiple linear regression analysis^12^ was then performed for confirmed cases at the national level using estimated populations and mean maximum temperatures at assigned latitudes as predictors. Case data was transformed using the Box-Cox^14^ method with λ = 0. To reduce variance, populations were then binned^11^ into 5° latitude and temperature ranges set at 2.5° intervals. Multiple linear regression was then performed for confirmed cases binned by latitude, again using population and mean maximum temperature as predictors, with and without a categorical variable describing bins above and below 30° latitude. Confirmed case data binned by latitude was transformed, again using the Box-Cox method, with λ = 0.212028 for analysis including the categorical variable and λ = 0 for analysis without. Nonlinear regression analysis^13^ was also performed, without transformation, for case rates binned by mean maximum temperature at latitude, using temperature as the only predictor.

No other adjustments were made to original data. Inclusion of multiple dates and binning of data is intended to reduce low-level (i.e. national level) variance associated with local weather, travel restrictions, business closures and other countermeasures. No speculation is made outside the scope of the data.

## Results and Discussion

***Confirmed cases* and *case rates* plotted by *latitude* (fig. 3)** showed a separation of cases in nations with central or southern latitudes north of 30°. As of March 27, case rates also appeared to be increasing south of -30° latitude, where temperatures fall first with transition to the fall season. That increase provided initial indication that the correlation with latitude is likely due to the underlying relationship with temperature.

**Figure 3:**
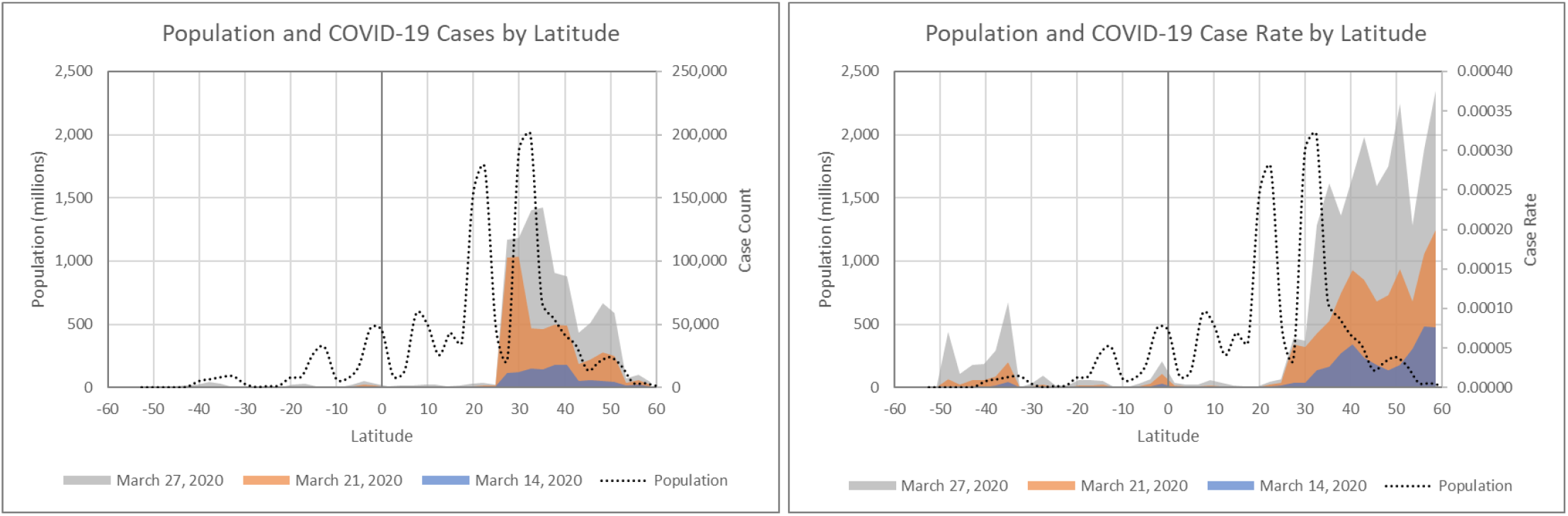
Confirmed cases and case rates by latitude for March 14, 21 and 27, 2020. The separation between cases above and below 30° latitude is clearly visible. Case rates also appear to increase below -30° latitude, where temperatures are lowest in the southern hemisphere.

**Binned *confirmed cases* and *case rates* plotted by *temperature* (fig. 4)** showed increased growth patterns in ranges below 22.5°C, and a downward trend could be seen in case rates as temperatures increase to the same point. Above 22.5°C, case rates remained near-zero. It should also be noted that high-growth regions appeared to be warming between March 14 and 27, but case rates remained near-zero in regions above 22.5°C during that time. Slight growth was observed above that temperature, but rates are significantly slower.

**Figure 4:**
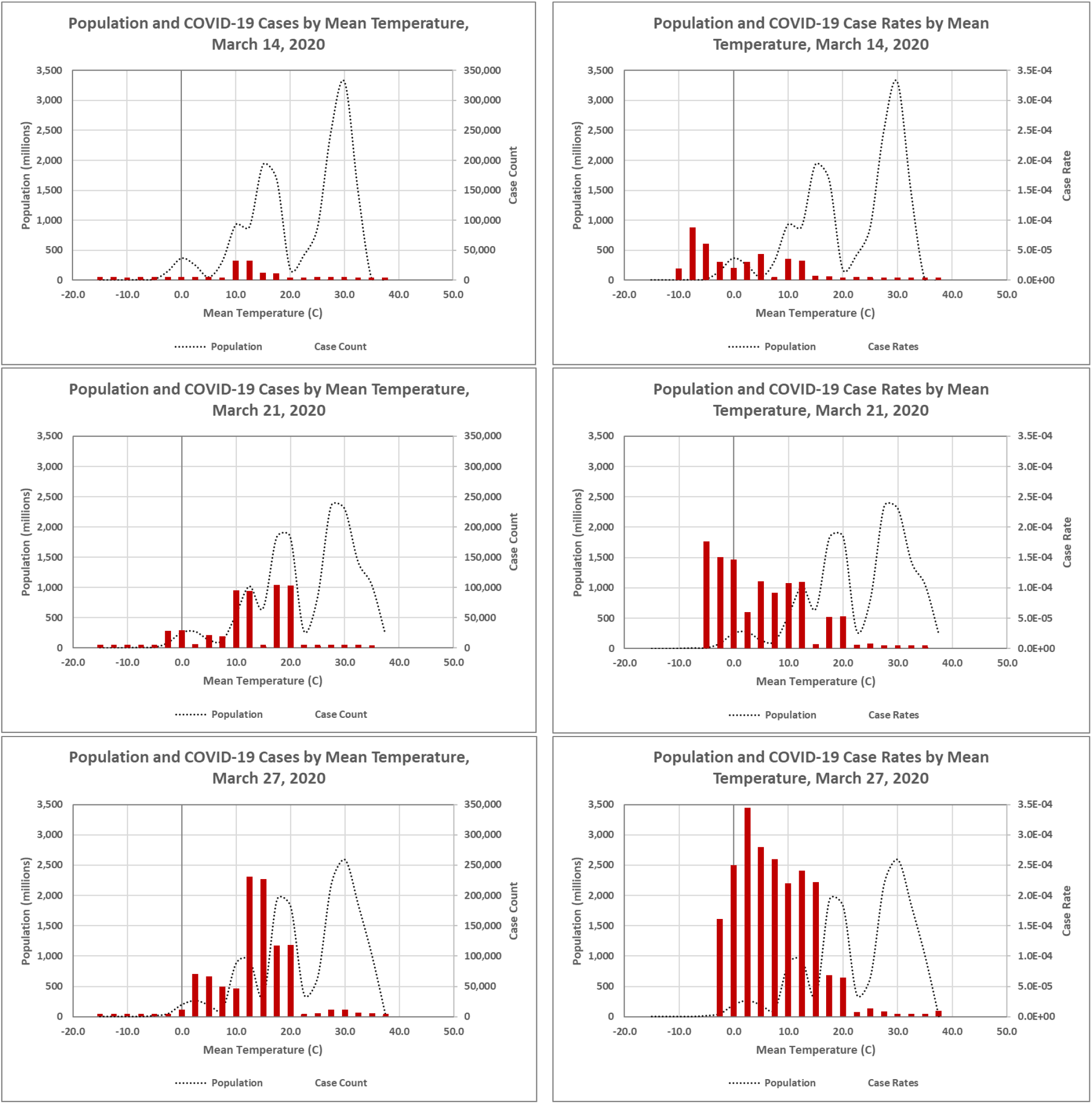
Confirmed cases and case rates by mean maximum temperature for March 14, March 21 and March 27, 2020. The trend of decreasing case rates with increased temperature, until approximately 22.5°C, above which case rates remain near-zero.

**Multiple linear regression of *confirmed cases* by *population* and *temperature***, for data reported March 27, 2020, showed a significant relationship between population and mean maximum temperature at the assigned latitude, with reported p-values <0.000 for both predictors and the constant. Residual values and distribution also indicated a good fit for the model. However, R^2^ of 26.52% showed that the model could only explain a small portion of low-level variance.

**Multiple linear regression of binned *confirmed cases* by *population, temperature* and *latitude (above/below 30*** °) also showed a significant relationship between response and continuous predictors. Transformed R^2^ improved to 84.61%, but the categorical variable for cases above and below 30° latitude could only be claimed with 84.7% confidence. Removing that variable showed similar significance for temperature and population predictors but reduced transformed R^2^ to 65.27%. R^2^ only improved to 30.92%, which is still an insignificant amount of variance explained. Normal probability plots of residuals also indicate a poor fit for the model – normality was rejected with a p-value of 0.028.

**Table 3:** Summary of nonlinear regression results. R^2^ of 93.78% shows that the model can explain the majority of binned variance.

| Nonlinear Regression Summary |
| --- |
| Case Rate by Temperature = $(0.0002166 - 7.03186\text{e-}006 \text{ Temperature}) / (1 - 0.0930346 \text{ Temperature} + 0.00495066 * \text{Temperature}^2)$ |
| R<sup>2</sup> = 93.78% |

**Figure X:**
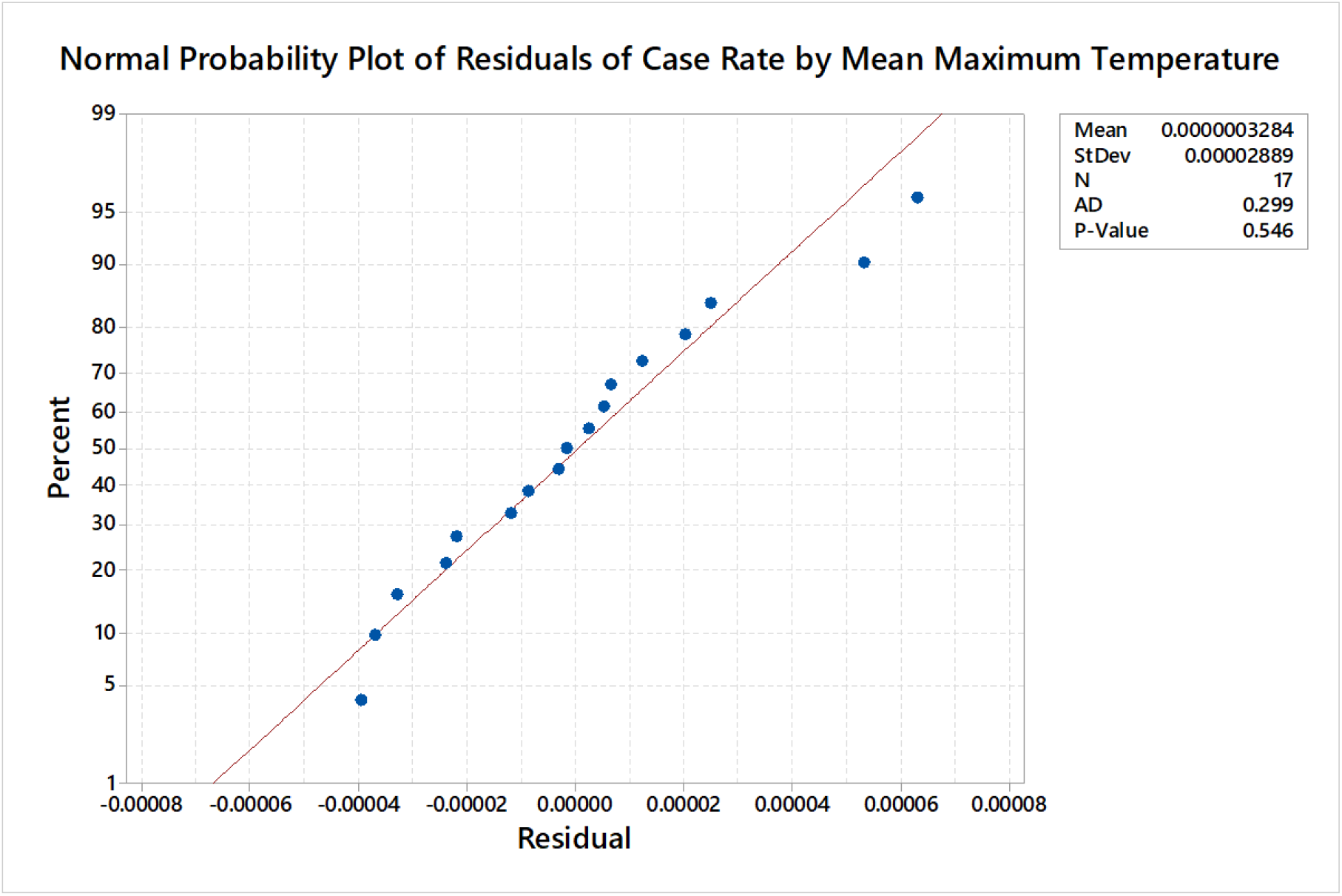
Normal probability of residuals for *case rates* by *mean maximum temperature*. The normal distribution of residuals with mean near-zero indicate a good fit for the model.

**Nonlinear regression of *case rates* by *temperature*** returned the best fit of all presented models. P-values cannot be calculated for independent nonlinear regression variables, so significance could not be determined with that method. However, R^2^ of 93.78% and normality test of residuals with p-value = 0.546 indicated a good fit for the model until case rates converge near zero above 22.5°C. Fig. 7 shows the close fit between predicted and actual values in that range.

**Table 1:**
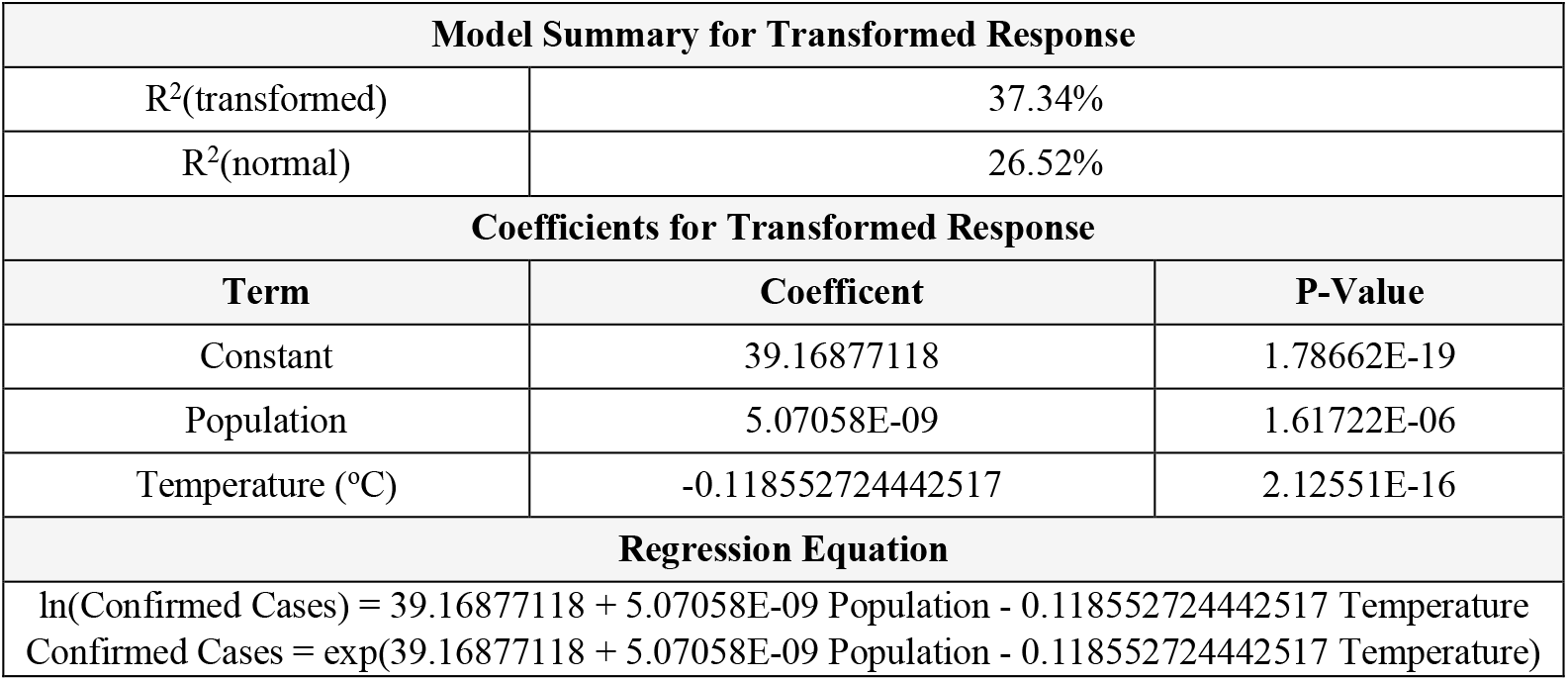
Summary of multiple linear regression of confirmed cases by population and temperature. P-values indicate statistical significance of predictors, but R^2^ indicates that the model explains only a small portion of low-level variance.

**Figure 5:**
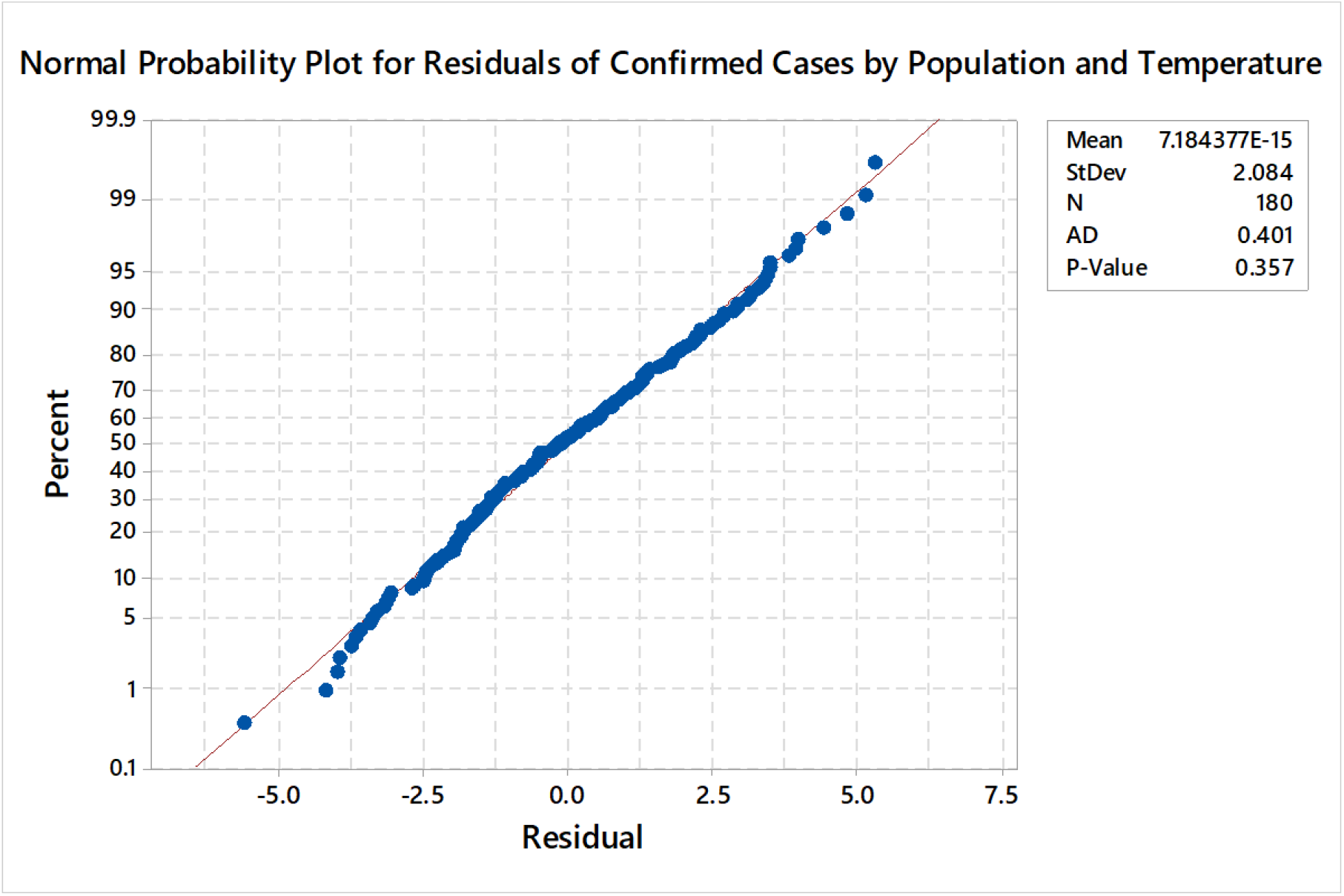
Normal probability plot for residuals of confirmed cases at the national level, as predicted by population and temperature. The normal distribution of residuals and mean near zero indicate a good fit for the model.

**Table 2:** Summary of multiple linear regression results. P-values indicate statistical significance of predictors, but R^2^ indicates that the model explains only a small portion of binned variance.

| Model Summary for Transformed Response |  |  |
| --- | --- | --- |
| R<sup>2</sup>(transformed) | 65.27% |  |
| R<sup>2</sup>(normal) | 30.92% |  |
| Coefficients for Transformed Response |  |  |
| Term | Coefficient | P-Value |
| Constant | 10.58762191 | 1.68022E-21 |
| Population | -0.136589856 | 1.71103E-08 |
| Temperature (°C) | 1.77677E-09 | 0.000379889 |
| Regression Equation |  |  |
| ln(Confirmed Cases) = 10.58762191 - 0.136589856 Population + 1.77677E-09 Temperature<br>Confirmed Cases = exp(10.58762191 - 0.136589856 Population + 1.77677E-09 Temperature ) |  |  |

**Figure 6:**
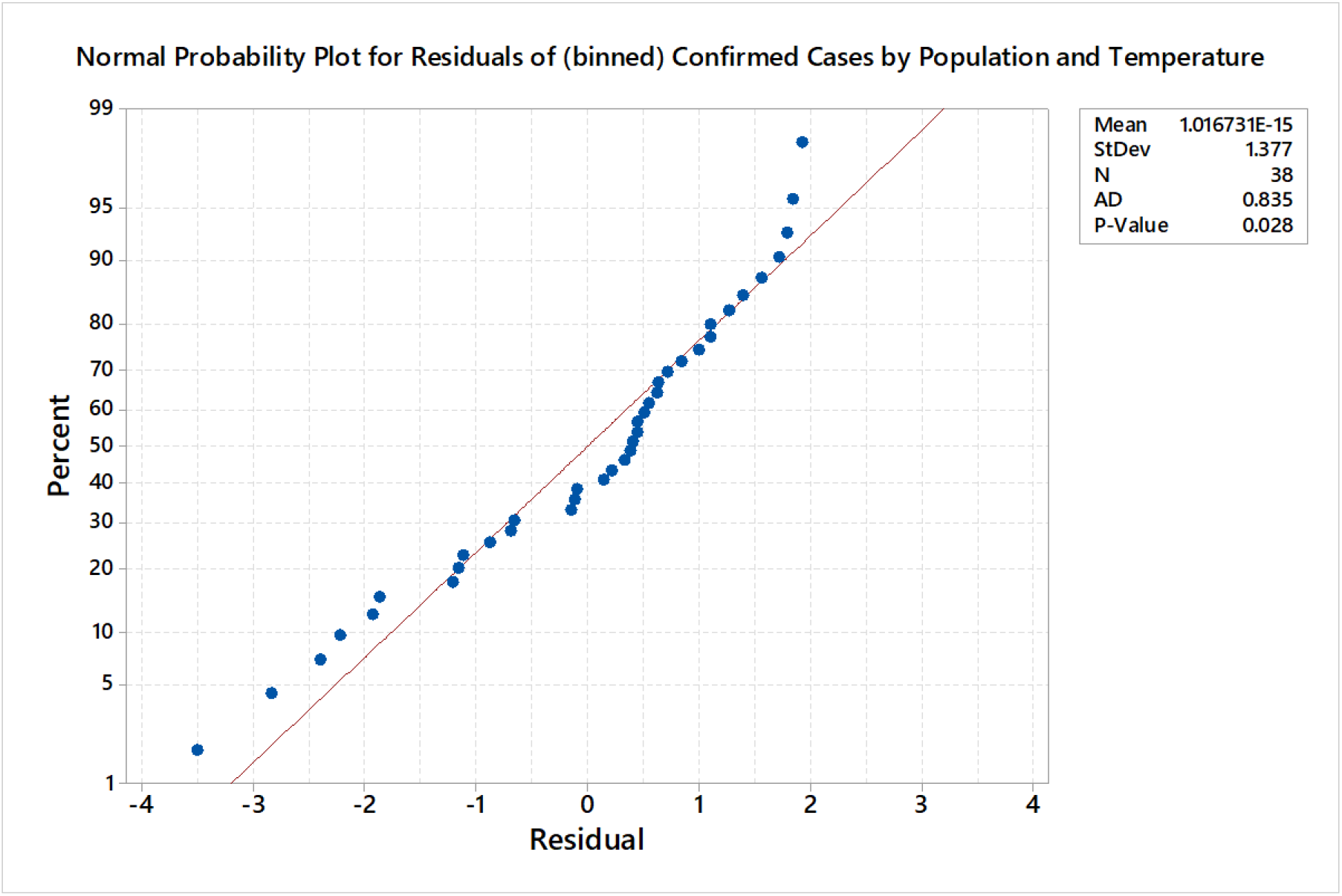
Normal probability plot for residuals of (binned) confirmed cases by population and temperature. The distribution of residuals indicates that the model is not a good fit for the mode.

**Figure 7:**
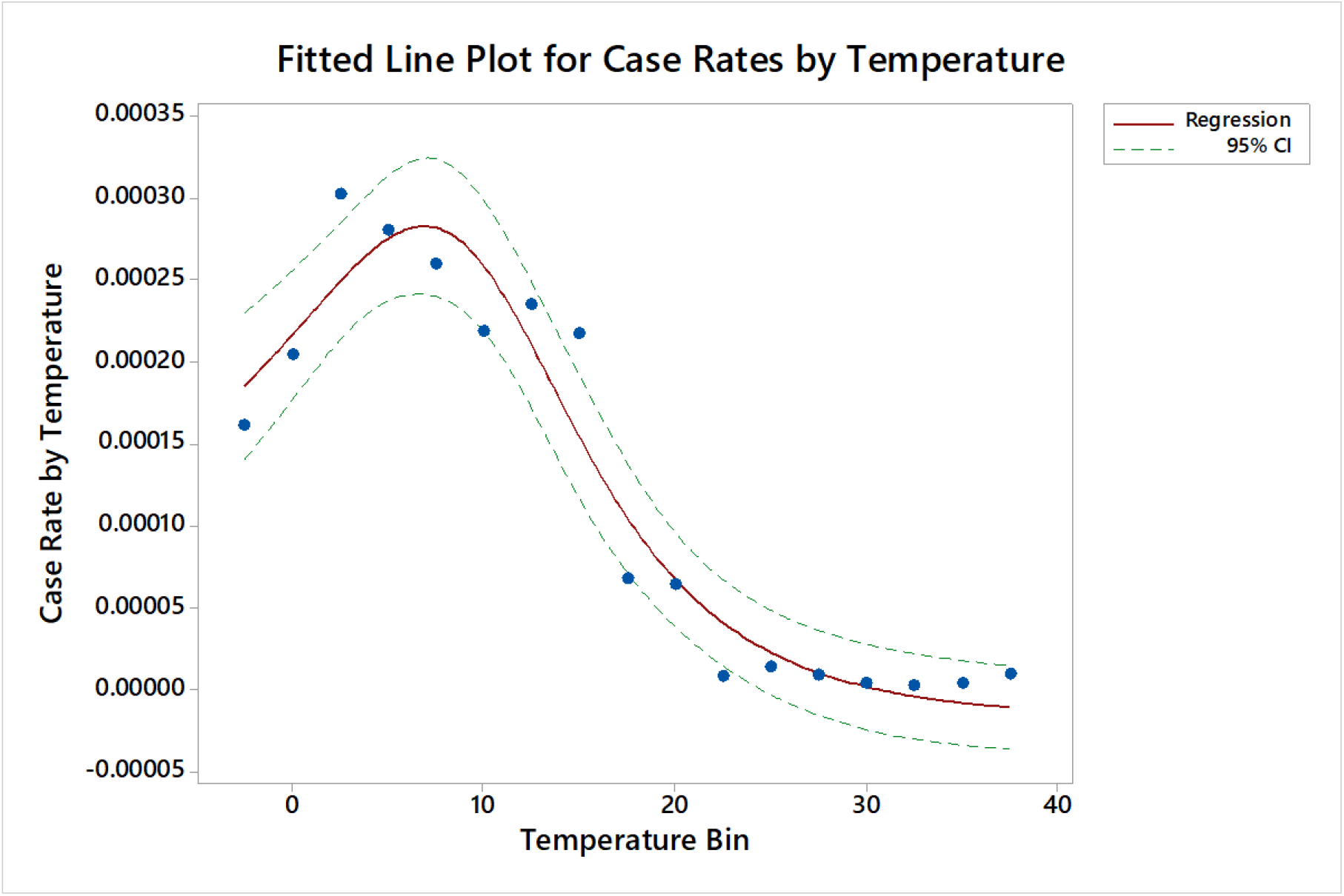
Fitted line plot for nonlinear regression of *case rates* by *temperature*. A close fit between predicted and actual values is apparent until the model begins to diverge from data (i.e. move below zero) above 22.5°C. Case rates remain near-zero above that temperature.

Modeling of case data reported March 27, 2020,as predicted by temperature, indicated a strong regional-level correlation between COVID-19 case rates and mean maximum surface air temperatures below 22.5°C. Case rates peaked in a goldilocks range around 7.5°C and were uniformly distributed near zero at temperatures above 22.5°C. That breakpoint was shown to be a consistent maximum across the dates included in analysis – i.e. case rates have been observed near zero below that temperature but have not been observed to increase significantly above it.

In conflict with the model for March 27, case rates reported for March 14 and 21 trended continuously upward with decreased temperature below 22.5°C. Nonlinearity for the most recent data might be explained by typical warming trends in northern latitudes that move case rate and population distributions to the right through March; variance in national- and local-level countermeasures may have effected growth in northern regions; the virus naturally peaks at lower levels in extremely cold and sparsely populated northern temperate/polar regions; and/or the virus was in a previous stage of global development. Regardless of cold weather dynamics, however, the breakpoint of 22.5°C remains apparent.

These conclusions do not confirm that COVID-19 cannot survive or transmit in warm and humid temperatures or establish a casual connection between temperature and transmission. However, the clear correlation between variables provides support for further study of SARS-CoV-2 and COVID-19 under various environmental conditions. With respect to countermeasures, the southern hemisphere should also expect increased case rates as that region moves from summer into fall and winter.

## Data Availability

Data is available for use by anyone for any purpose

## Appendix: Reported Data

| National Data |  |  | Average Temperature at Latitude |  |  | Confirmed Cases Reported |  |  |
| --- | --- | --- | --- | --- | --- | --- | --- | --- |
| Area | Study Latitude | Population | 14-Mar | 21-Mar | 27-Mar | 14-Mar | 21-Mar | 27-Mar |
| Afghanistan | 33.93 | 38,928,341 | 13.46 | 14.99 | 15.95 | 7 | 24 | 80 |
| Albania | 41.15 | 2,877,800 | 8.77 | 13.09 | 10.26 | 33 | 70 | 186 |
| Algeria | 28.13 | 43,851,043 | 22.06 | 23.94 | 25.14 | 26 | 94 | 305 |
| Angola | -11.21 | 32,866,268 | 26.37 | 25.41 | 25.96 | 0 | 0 | 2 |
| Antigua and Barbuda | 17.34 | 97,928 | 31.80 | 34.10 | 35.55 | 1 | 1 | 3 |
| Argentina | -38.48 | 45,195,777 | 23.48 | 20.78 | 20.23 | 34 | 128 | 502 |
| Armenia | 40.07 | 2,963,234 | 10.02 | 14.16 | 12.01 | 8 | 136 | 329 |
| Aruba | 12.49 | 106,766 | 34.59 | 36.02 | 36.85 | 0 | 5 | 19 |
| Australia | -32.21 | 25,499,881 | 27.28 | 26.10 | 22.68 | 197 | 873 | 2985 |
| Austria | 47.70 | 9,006,400 | 2.17 | 7.23 | 6.00 | 504 | 2649 | 7029 |
| Azerbaijan | 40.17 | 10,139,175 | 10.02 | 14.16 | 12.01 | 11 | 44 | 122 |
| Bahamas | 24.09 | 393,248 | 27.14 | 29.36 | 30.52 | 0 | 4 | 5 |
| Bahrain | 26.11 | 1,701,583 | 25.59 | 28.04 | 28.31 | 210 | 285 | 458 |
| Bangladesh | 23.51 | 164,689,383 | 27.44 | 29.65 | 30.84 | 3 | 17 | 48 |
| Barbados | 13.19 | 287,371 | 34.16 | 35.70 | 36.57 | 0 | 5 | 18 |
| Belarus | 53.71 | 9,449,321 | -0.38 | 1.85 | 2.78 | 21 | 57 | 86 |
| Belgium | 50.52 | 11,589,616 | 0.74 | 3.61 | 3.97 | 599 | 2257 | 6235 |
| Belize | 17.19 | 397,621 | 31.83 | 34.10 | 35.42 | 0 | 0 | 2 |
| Benin | 9.22 | 12,123,198 | 34.70 | 34.68 | 33.62 | 0 | 2 | 6 |
| Bermuda | 32.32 | 62,273 | 15.85 | 17.74 | 18.62 | 0 | 2 | 7 |
| Bhutan | 27.47 | 771,612 | 22.96 | 24.97 | 25.85 | 1 | 2 | 3 |
| Bolivia (Plurinational State of) | -16.28 | 11,673,029 | 29.31 | 29.28 | 31.99 | 3 | 16 | 39 |
| Bosnia and Herzegovina | 43.92 | 3,280,815 | 5.37 | 10.89 | 7.69 | 11 | 44 | 213 |
| Brazil | -14.29 | 212,559,409 | 29.02 | 27.85 | 28.43 | 98 | 621 | 2433 |
| Brunei Darussalam | 4.55 | 437,483 | 35.16 | 32.56 | 32.09 | 25 | 78 | 114 |
| Bulgaria | 42.73 | 6,948,445 | 6.13 | 11.47 | 8.20 | 7 | 127 | 264 |
| Burkina Faso | 12.25 | 20,903,278 | 34.77 | 36.02 | 36.86 | 2 | 40 | 146 |
| Cabo Verde | 16.00 | 555,988 | 32.24 | 34.59 | 35.67 | 0 | 1 | 3 |
| Cambodia | 12.08 | 16,718,971 | 34.77 | 36.02 | 36.86 | 7 | 51 | 98 |
| Cameroon | 7.37 | 26,545,864 | 34.85 | 34.23 | 32.89 | 2 | 22 | 75 |
| Canada | 41.68 | 37,742,157 | 7.93 | 12.34 | 9.30 | 176 | 846 | 3555 |
| Cayman Islands | 19.51 | 65,720 | 30.61 | 32.97 | 34.66 | 1 | 3 | 8 |
| Central African Republic | 6.61 | 4,829,764 | 34.70 | 33.17 | 32.23 | 0 | 1 | 5 |
| Chad | 15.47 | 16,425,859 | 32.07 | 34.70 | 35.41 | 0 | 1 | 5 |
| Chile | -37.11 | 19,116,209 | 22.63 | 19.99 | 19.19 | 43 | 434 | 1306 |
| China | 31.20 | 1,439,323,774 | 18.00 | 19.89 | 21.01 | 0 | 81416 | 82078 |
| Colombia | 5.91 | 50,882,884 | 35.13 | 32.99 | 32.30 | 16 | 145 | 470 |
| Congo | -0.72 | 5,518,092 | 31.36 | 30.16 | 30.70 | 0 | 3 | 4 |
| Costa Rica | 8.28 | 5,094,114 | 34.91 | 34.83 | 33.57 | 23 | 113 | 201 |
| Côte d'Ivoire | 7.45 | 26,378,275 | 34.85 | 34.23 | 32.89 | 0 | 9 | 80 |
| Croatia | 44.37 | 4,105,268 | 4.94 | 10.52 | 7.59 | 27 | 126 | 495 |
| Cuba | 21.55 | 11,326,616 | 28.73 | 31.18 | 32.60 | 4 | 16 | 67 |
| Curaçao | 12.17 | 164,100 | 34.77 | 36.02 | 36.86 | 0 | 3 | 7 |
| Cyprus | 35.18 | 1,207,361 | 12.20 | 14.22 | 14.36 | 14 | 67 | 146 |
| Czechia | 49.80 | 10,708,982 | 0.86 | 4.15 | 4.27 | 150 | 904 | 2062 |
| Democratic Republic of the Congo | -4.03 | 89,561,404 | 30.00 | 29.62 | 30.45 | 2 | 14 | 54 |
| Denmark | 56.20 | 5,792,203 | -2.40 | 0.89 | 2.16 | 801 | 1255 | 1877 |
| Djibouti | 11.85 | 988,002 | 34.77 | 36.02 | 36.86 | 0 | 1 | 12 |
| Dominica | 15.40 | 71,991 | 32.07 | 34.70 | 35.41 | 0 | 0 | 11 |
| Dominican Republic | 19.29 | 10,847,904 | 30.61 | 32.97 | 34.66 | 5 | 72 | 488 |
| Ecuador | -1.57 | 17,643,060 | 30.73 | 29.73 | 30.84 | 23 | 367 | 1211 |
| Egypt | 26.92 | 102,334,403 | 23.98 | 26.07 | 26.82 | 93 | 256 | 495 |
| El Salvador | 13.71 | 6,486,201 | 33.63 | 35.43 | 36.17 | 0 | 1 | 13 |
| Equatorial Guinea | 1.16 | 1,402,985 | 32.37 | 30.98 | 30.35 | 0 | 4 | 6 |
| Eritrea | 15.21 | 3,546,427 | 31.90 | 34.52 | 35.03 | 0 | 0 | 6 |
| Estonia | 58.72 | 1,326,539 | -3.85 | 0.22 | 1.97 | 79 | 283 | 538 |
| Eswatini | 26.52 | 1,160,164 | 24.97 | 27.33 | 27.81 | 0 | 1 | 6 |
| Ethiopia | 9.15 | 114,963,583 | 34.70 | 34.68 | 33.62 | 1 | 9 | 12 |
| Fiji | -17.10 | 896,444 | 29.47 | 29.07 | 32.04 | 0 | 1 | 5 |
| Finland | 59.45 | 5,540,718 | -5.04 | -0.76 | 1.11 | 109 | 450 | 958 |
| France | 46.27 | 65,273,512 | 2.94 | 8.46 | 6.59 | 3640 | 12475 | 28786 |
| French Guiana | 3.93 | 298,682 | 34.72 | 32.13 | 31.68 | 6 | 15 | 28 |
| French Polynesia | -17.88 | 280,904 | 29.64 | 28.74 | 31.97 | 1 | 11 | 30 |
| Gabon | -0.89 | 2,225,728 | 31.09 | 30.05 | 30.80 | 1 | 3 | 6 |
| Gambia | 13.44 | 2,416,664 | 33.63 | 35.43 | 36.17 | 0 | 1 | 2 |
| Georgia | 42.32 | 3,989,175 | 6.13 | 11.47 | 8.20 | 25 | 43 | 81 |
| Germany | 51.18 | 83,783,945 | 0.69 | 3.28 | 3.85 | 3062 | 18323 | 42288 |
| Ghana | 7.86 | 31,072,945 | 34.96 | 34.68 | 33.39 | 1 | 16 | 132 |
| Greece | 38.22 | 10,423,056 | 10.87 | 13.75 | 13.98 | 98 | 495 | 892 |
| Greenland | 59.52 | 56,772 | -5.04 | -0.76 | 1.11 | 0 | 2 | 6 |
| Grenada | 12.19 | 112,519 | 34.77 | 36.02 | 36.86 | 0 | 0 | 7 |
| Guadeloupe | 16.17 | 400,127 | 32.24 | 34.59 | 35.67 | 1 | 51 | 76 |
| Guam | 13.44 | 168,783 | 33.63 | 35.43 | 36.17 | 0 | 14 | 45 |
| Guatemala | 15.73 | 17,915,567 | 32.07 | 34.70 | 35.41 | 0 | 12 | 24 |
| Guinea | 9.93 | 13,132,792 | 34.48 | 34.96 | 34.75 | 1 | 2 | 5 |
| Guyana | 4.89 | 786,559 | 35.25 | 32.83 | 32.25 | 1 | 5 | 5 |
| Haiti | 19.06 | 11,402,533 | 30.85 | 33.30 | 35.01 | 0 | 2 | 8 |
| Honduras | 15.30 | 9,904,608 | 32.07 | 34.70 | 35.41 | 2 | 24 | 52 |
| Hungary | 47.16 | 9,660,350 | 2.45 | 7.78 | 6.21 | 19 | 85 | 300 |
| India | 21.11 | 1,380,004,385 | 29.26 | 31.99 | 33.21 | 82 | 195 | 724 |
| Indonesia | -2.47 | 273,523,621 | 30.42 | 29.45 | 30.81 | 69 | 369 | 893 |
| Iran (Islamic Republic of) | 32.31 | 83,992,953 | 15.85 | 17.74 | 18.62 | 11364 | 19644 | 29406 |
| Iraq | 33.22 | 40,222,503 | 14.88 | 16.63 | 17.69 | 93 | 193 | 382 |
| Ireland | 53.43 | 4,937,796 | -0.38 | 1.85 | 2.78 | 90 | 683 | 1819 |
| Isle of Man | 54.24 | 85,032 | -0.82 | 1.43 | 2.36 | 0 | 1 | 26 |
| Israel | 31.39 | 8,655,541 | 17.51 | 19.56 | 20.41 | 100 | 712 | 3035 |
| Italy | 41.19 | 60,461,828 | 8.77 | 13.09 | 10.26 | 17660 | 47021 | 80539 |
| Jamaica | 17.66 | 2,961,161 | 31.80 | 34.10 | 35.55 | 7 | 16 | 26 |
| Japan | 32.96 | 126,476,458 | 14.88 | 16.63 | 17.69 | 716 | 996 | 1387 |
| Jordan | 31.28 | 10,203,140 | 17.51 | 19.56 | 20.41 | 1 | 69 | 212 |
| Kazakhstan | 48.01 | 18,776,707 | 1.82 | 6.53 | 5.68 | 0 | 53 | 125 |
| Kenya | -0.14 | 53,771,300 | 31.76 | 30.53 | 30.59 | 1 | 7 | 25 |
| Kuwait | 29.31 | 4,270,563 | 19.55 | 21.51 | 22.82 | 100 | 159 | 208 |
| Kyrgyzstan | 41.22 | 6,524,191 | 8.77 | 13.09 | 10.26 | 0 | 6 | 58 |
| Lao People's Democratic Republic | 18.21 | 7,275,556 | 31.49 | 33.83 | 35.40 | 0 | 0 | 6 |
| Latvia | 56.88 | 1,886,202 | -2.92 | 0.74 | 2.38 | 16 | 111 | 244 |
| Lebanon | 33.87 | 6,825,442 | 13.46 | 14.99 | 15.95 | 77 | 163 | 368 |
| Liberia | 6.35 | 5,057,677 | 34.70 | 33.17 | 32.23 | 0 | 2 | 3 |
| Libya | 26.43 | 6,871,287 | 24.97 | 27.33 | 27.81 | 0 | 0 | 1 |
| Lithuania | 55.17 | 2,722,291 | -1.47 | 0.96 | 1.91 | 6 | 69 | 299 |
| Madagascar | -18.78 | 27,691,019 | 29.90 | 28.86 | 32.13 | 0 | 0 | 24 |
| Malaysia | 2.39 | 32,365,998 | 33.39 | 31.43 | 30.83 | 197 | 1030 | 2031 |
| Maldives | 3.20 | 540,542 | 33.99 | 31.75 | 31.13 | 9 | 13 | 13 |
| Malta | 35.94 | 441,539 | 12.01 | 14.34 | 14.17 | 12 | 64 | 134 |
| Martinique | 14.64 | 375,265 | 32.36 | 34.74 | 35.30 | 6 | 32 | 66 |
| Mauritania | 21.02 | 4,649,660 | 29.26 | 31.99 | 33.21 | 0 | 2 | 3 |
| Mauritius | -15.43 | 1,271,767 | 28.68 | 28.76 | 29.99 | 0 | 12 | 81 |
| Mayotte | -12.83 | 272,813 | 28.23 | 26.51 | 26.80 | 0 | 4 | 50 |
| Mexico | 23.55 | 128,932,753 | 27.44 | 29.65 | 30.84 | 26 | 164 | 478 |
| Monaco | 43.74 | 39,244 | 5.78 | 11.25 | 7.88 | 2 | 12 | 19 |
| Mongolia | 46.86 | 3,278,292 | 2.45 | 7.78 | 6.21 | 1 | 6 | 11 |
| Montenegro | 42.65 | 628,062 | 6.13 | 11.47 | 8.20 | 0 | 14 | 67 |
| Montserrat | 16.75 | 4,999 | 31.98 | 34.36 | 35.64 | 0 | 1 | 2 |
| Morocco | 28.69 | 36,910,558 | 20.92 | 22.95 | 24.27 | 7 | 74 | 275 |
| Mozambique | -18.62 | 31,255,435 | 29.82 | 28.66 | 31.87 | 0 | 0 | 5 |
| Myanmar | 18.99 | 54,409,794 | 30.85 | 33.30 | 35.01 | 0 | 0 | 5 |
| Namibia | -22.97 | 2,540,916 | 32.90 | 34.02 | 34.88 | 0 | 3 | 8 |
| Nepal | 28.40 | 29,136,808 | 20.92 | 22.95 | 24.27 | 1 | 1 | 3 |
| Netherlands | 52.23 | 17,134,873 | 0.60 | 3.08 | 3.63 | 804 | 2994 | 7431 |
| New Caledonia | -20.45 | 285,491 | 30.87 | 30.92 | 33.39 | 0 | 2 | 14 |
| New Zealand | -40.93 | 4,822,233 | 22.45 | 20.87 | 19.14 | 6 | 53 | 338 |
| Nicaragua | 12.87 | 6,624,554 | 34.16 | 35.70 | 36.57 | 0 | 1 | 2 |
| Niger | 17.61 | 24,206,636 | 31.80 | 34.10 | 35.55 | 0 | 1 | 10 |
| Nigeria | 8.98 | 206,139,587 | 34.70 | 34.68 | 33.62 | 2 | 12 | 65 |
| North Macedonia | 41.61 | 2,083,380 | 7.93 | 12.34 | 9.30 | 9 | 70 | 201 |
| Norway | 57.76 | 5,421,242 | -3.65 | 0.43 | 2.06 | 750 | 1742 | 3156 |
| Oman | 21.58 | 5,106,622 | 28.73 | 31.18 | 32.60 | 19 | 48 | 109 |
| Pakistan | 30.31 | 220,892,331 | 18.59 | 20.31 | 21.58 | 21 | 461 | 1057 |
| Panama | 8.45 | 4,314,768 | 34.91 | 34.83 | 33.57 | 27 | 137 | 558 |
| Papua New Guinea | -5.68 | 8,947,027 | 30.10 | 29.79 | 30.95 | 0 | 1 | 1 |
| Paraguay | -23.45 | 7,132,530 | 33.09 | 34.04 | 34.60 | 6 | 13 | 41 |
| Peru | -10.12 | 32,971,846 | 27.29 | 26.02 | 27.09 | 28 | 234 | 580 |
| Philippines | 12.77 | 109,581,085 | 34.16 | 35.70 | 36.57 | 64 | 230 | 707 |
| Poland | 52.02 | 37,846,605 | 0.60 | 3.08 | 3.63 | 64 | 425 | 1221 |
| Portugal | 35.99 | 10,196,707 | 12.01 | 14.34 | 14.17 | 112 | 1020 | 3544 |
| Puerto Rico | 18.22 | 2,860,840 | 31.49 | 33.83 | 35.40 | 3 | 14 | 64 |
| Qatar | 25.43 | 2,881,060 | 25.79 | 28.14 | 28.35 | 262 | 460 | 549 |
| Republic of Korea | 35.77 | 51,269,183 | 12.01 | 14.34 | 14.17 | 8086 | 8799 | 9332 |
| Republic of Moldova | 46.98 | 4,033,963 | 2.45 | 7.78 | 6.21 | 8 | 66 | 177 |
| Réunion | -21.13 | 895,308 | 31.55 | 31.98 | 34.19 | 5 | 28 | 135 |
| Romania | 45.94 | 19,237,682 | 3.49 | 8.97 | 7.06 | 64 | 308 | 1029 |
| Russian Federation | 41.19 | 145,934,460 | 8.77 | 13.09 | 10.26 | 34 | 253 | 1036 |
| Rwanda | -1.94 | 12,952,209 | 30.42 | 29.47 | 30.77 | 0 | 11 | 50 |
| Saint Barthélemy | 17.90 | 9,885 | 31.49 | 33.83 | 35.40 | 0 | 3 | 3 |
| Saint Lucia | 13.89 | 183,629 | 32.89 | 35.04 | 35.50 | 0 | 2 | 3 |
| Saint Vincent and the Grenadines | 13.05 | 110,947 | 34.16 | 35.70 | 36.57 | 1 | 1 | 1 |
| Saudi Arabia | 24.22 | 34,813,867 | 27.14 | 29.36 | 30.52 | 62 | 274 | 1012 |
| Senegal | 14.46 | 16,743,930 | 32.36 | 34.74 | 35.30 | 10 | 38 | 105 |
| Serbia | 44.21 | 8,737,370 | 5.37 | 10.89 | 7.69 | 31 | 135 | 457 |
| Seychelles | -6.99 | 98,340 | 29.60 | 29.55 | 30.17 | 0 | 6 | 7 |
| Singapore | 1.29 | 5,850,343 | 32.70 | 31.20 | 30.45 | 200 | 385 | 683 |
| Slovakia | 48.67 | 5,459,643 | 1.47 | 6.02 | 5.27 | 30 | 137 | 226 |
| Slovenia | 46.15 | 2,078,932 | 3.49 | 8.97 | 7.06 | 141 | 341 | 577 |
| Somalia | 5.19 | 15,893,219 | 35.25 | 32.83 | 32.25 | 0 | 1 | 3 |
| South Africa | -34.65 | 59,308,690 | 25.75 | 24.08 | 22.20 | 17 | 205 | 927 |
| Spain | 35.71 | 46,754,783 | 11.76 | 14.11 | 13.76 | 4231 | 19980 | 56188 |
| Sri Lanka | 7.88 | 21,413,250 | 34.96 | 34.68 | 33.39 | 6 | 59 | 106 |
| Sudan | 15.46 | 43,849,269 | 32.07 | 34.70 | 35.41 | 1 | 1 | 3 |
| Suriname | 4.03 | 586,634 | 34.72 | 32.13 | 31.68 | 0 | 2 | 7 |
| Sweden | 55.13 | 10,099,270 | -1.47 | 0.96 | 1.91 | 775 | 1623 | 2806 |
| Switzerland | 46.81 | 8,654,618 | 2.45 | 7.78 | 6.21 | 1125 | 4840 | 10714 |
| Syrian Arab Republic | 34.81 | 17,500,657 | 12.20 | 14.22 | 14.36 | 0 | 0 | 5 |
| Thailand | 13.04 | 69,799,978 | 34.16 | 35.70 | 36.57 | 75 | 322 | 1136 |
| Timor-Leste | -8.83 | 1,318,442 | 28.10 | 27.18 | 28.02 | 0 | 1 | 1 |
| Togo | 8.53 | 8,278,737 | 34.91 | 34.83 | 33.57 | 1 | 9 | 24 |
| Trinidad and Tobago | 10.72 | 1,399,491 | 34.39 | 35.11 | 35.30 | 1 | 9 | 61 |
| Tunisia | 34.00 | 11,818,618 | 13.46 | 14.99 | 15.95 | 16 | 54 | 197 |
| Turkey | 39.05 | 84,339,067 | 10.40 | 13.97 | 12.67 | 5 | 670 | 3629 |
| Turks and Caicos Islands | 21.56 | 38,718 | 28.73 | 31.18 | 32.60 | 0 | 0 | 2 |
| Uganda | 1.38 | 45,741,000 | 32.70 | 31.20 | 30.45 | 0 | 0 | 14 |
| Ukraine | 48.28 | 43733759 | 1.47 | 6.02 | 5.27 | 3 | 26 | 218 |
| United Arab Emirates | 24.46 | 9890400 | 26.59 | 28.73 | 29.86 | 85 | 140 | 333 |
| United Republic of Tanzania | -6.37 | 59734213 | 29.64 | 29.40 | 30.29 | 0 | 6 | 13 |
| United States of America | 37.26 | 331002647 | 11.08 | 13.79 | 14.42 | 1678 | 15219 | 68334 |
| United States Virgin Islands | 18.04 | 104423 | 31.49 | 33.83 | 35.40 | 0 | 3 | 17 |
| Uruguay | -32.93 | 3473727 | 26.28 | 25.26 | 21.58 | 0 | 94 | 217 |
| Uzbekistan | 41.38 | 33,469,199 | 7.93 | 12.34 | 9.30 | 0 | 33 | 83 |
| Venezuela (Bolivarian Republic of) | 8.28 | 28,435,943 | 34.91 | 34.83 | 33.57 | 2 | 36 | 91 |
| Viet Nam | 15.78 | 97,338,583 | 32.24 | 34.59 | 35.67 | 48 | 91 | 153 |
| Zambia | -13.1 | 18,383,956 | 28.23 | 26.51 | 26.80 | 0 | 2 | 3 |
| Zimbabwe | -19.01 | 14,862,927 | 29.90 | 28.86 | 32.13 | 0 | 1 | 3 |

| Binned Latitude Data |  | Average Temperature at Latitude |  |  | Confirmed Cases near Latitude (+/- 2.5°) |  |  |
| --- | --- | --- | --- | --- | --- | --- | --- |
| Study Latitude | Population | 14-Mar | 21-Mar | 27-Mar | 14-Mar | 21-Mar | 27-Mar |
| -42.50 | 4,822,233 | 17.0514 | 15.5040 | 13.3142 | 6 | 53 | 338 |
| -40.00 | 50,018,010 | 20.4288 | 20.9089 | 20.0613 | 40 | 181 | 840 |
| -37.50 | 64,311,986 | 22.3444 | 18.9643 | 18.8127 | 77 | 562 | 1808 |
| -35.00 | 81,898,626 | 24.2398 | 23.6912 | 21.5318 | 60 | 733 | 2450 |
| -32.50 | 88,282,298 | 25.7765 | 25.7108 | 22.0398 | 214 | 1172 | 4129 |
| -30.00 | 27,642,133 | 27.3579 | 27.4348 | 25.4497 | 197 | 873 | 2985 |
| -27.50 | 2,142,252 | 31.4486 | 30.9216 | 29.4972 | 0 | 0 | 0 |
| -25.00 | 9,673,446 | 32.8217 | 33.1379 | 32.9120 | 6 | 16 | 49 |
| -22.50 | 13,205,870 | 33.8631 | 33.9473 | 35.1159 | 11 | 46 | 198 |
| -20.00 | 77,730,024 | 32.8355 | 30.0532 | 32.9469 | 6 | 42 | 211 |
| -17.50 | 88,363,554 | 32.1087 | 28.8910 | 32.0003 | 4 | 41 | 187 |
| -15.00 | 264,765,694 | 29.4980 | 28.1486 | 28.8868 | 101 | 656 | 2611 |
| -12.50 | 317,307,449 | 27.3179 | 26.4357 | 26.7356 | 126 | 861 | 3068 |
| -10.00 | 68,726,171 | 26.8855 | 26.0163 | 27.0934 | 28 | 235 | 583 |
| -7.50 | 70,798,042 | 30.1384 | 29.4007 | 30.0222 | 0 | 14 | 22 |
| -5.00 | 170,231,765 | 30.4975 | 30.0010 | 30.9690 | 2 | 27 | 75 |
| -2.50 | 467,097,029 | 30.9723 | 29.4500 | 30.8117 | 96 | 774 | 2243 |
| 0.00 | 451,343,777 | 32.4381 | 30.5346 | 30.5898 | 491 | 2179 | 4923 |
| 2.50 | 88,348,833 | 33.3472 | 31.4346 | 30.8308 | 438 | 1532 | 2901 |
| 5.00 | 132,314,869 | 34.8597 | 32.8335 | 32.2464 | 59 | 293 | 803 |
| 7.50 | 582,610,871 | 33.5747 | 34.2346 | 32.8859 | 82 | 575 | 1836 |
| 10.00 | 495,307,027 | 32.9475 | 34.9605 | 34.7508 | 74 | 504 | 1550 |
| 12.50 | 255,139,532 | 32.9600 | 36.0233 | 36.8464 | 166 | 756 | 2448 |
| 15.00 | 433,224,091 | 31.2306 | 34.5190 | 35.0264 | 209 | 830 | 2438 |
| 17.50 | 354,855,972 | 31.0251 | 34.1049 | 35.5537 | 69 | 297 | 975 |
| 20.00 | 1,535,536,289 | 29.7731 | 32.7902 | 34.3128 | 121 | 375 | 1540 |
| 22.50 | 1,740,442,982 | 28.2798 | 30.1638 | 31.4773 | 281 | 860 | 2781 |
| 25.00 | 455,037,090 | 25.4406 | 28.2626 | 28.9025 | 742 | 1603 | 3388 |
| 27.50 | 229,889,081 | 22.6477 | 24.9666 | 25.8471 | 700 | 1332 | 2303 |
| 30.00 | 1,877,298,984 | 19.6853 | 20.8464 | 22.0935 | 11620 | 102632 | 116586 |
| 32.50 | 2,004,902,031 | 17.2600 | 17.7423 | 18.6180 | 12395 | 103734 | 118214 |
| 35.00 | 682,677,930 | 13.0539 | 14.2248 | 14.3632 | 15042 | 46579 | 140097 |
| 37.50 | 551,236,863 | 11.8287 | 13.7874 | 14.4242 | 14236 | 46314 | 142199 |
| 40.00 | 416,516,360 | 12.2343 | 14.1597 | 12.0144 | 18059 | 49687 | 90711 |
| 42.50 | 330,035,815 | 9.0442 | 11.4685 | 8.2018 | 18034 | 48980 | 87705 |
| 45.00 | 136,111,688 | 8.4701 | 10.0423 | 7.4376 | 5076 | 18579 | 43109 |
| 47.50 | 200,572,748 | 3.9762 | 7.2327 | 5.9971 | 5685 | 21890 | 51254 |
| 50.00 | 238,666,506 | 1.6644 | 4.1517 | 4.2704 | 5216 | 27768 | 66835 |
| 52.50 | 164,827,188 | 0.2501 | 2.7511 | 3.4056 | 4640 | 24740 | 59106 |
| 55.00 | 34,972,115 | -2.1001 | 0.9623 | 1.9090 | 1709 | 3799 | 7157 |
| 57.50 | 32,845,237 | -2.7305 | 0.5839 | 2.2285 | 2536 | 5535 | 9884 |
| 60.00 | 12,394,136 | -4.9678 | -1.0675 | 0.5952 | 938 | 2477 | 4658 |

| Binned Temperature Data | Population |  |  | Confirmed Cases |  |  |
| --- | --- | --- | --- | --- | --- | --- |
| Study Temperature (deg C) | 14-Mar | 21-Mar | 27-Mar | 14-Mar | 21-Mar | 27-Mar |
| -10 | 341,250 | 0 | 0 | 109 | 0 | 0 |
| -7.5 | 390,115 | 0 | 0 | 1,755 | 0 | 0 |
| -5 | 14,280,338 | 390,115 | 341,250 | 2,538 | 2,588 | 0 |
| -2.5 | 47,317,386 | 5,987,605 | 390,115 | 5,421 | 28,018 | 964 |
| 0 | 194,775,910 | 47,317,386 | 32,979,134 | 4,712 | 28,591 | 11,815 |
| 2.5 | 264,331,866 | 203,409,893 | 209,007,383 | 5,480 | 6,026 | 70,088 |
| 5 | 189,237,790 | 238,666,506 | 365,314,904 | 5,529 | 20,986 | 66,835 |
| 7.5 | 114,436,312 | 189,237,790 | 290,372,917 | 493 | 19,571 | 49,192 |
| 10 | 880,065,317 | 213,396,408 | 330,035,815 | 32,243 | 95,227 | 47,029 |
| 12.5 | 928,505,345 | 956,345,736 | 797,638,208 | 32,891 | 94,085 | 230,554 |
| 15 | 327,034,606 | 1,021,909,570 | 626,309,921 | 12,374 | 1,931 | 226,888 |
| 17.5 | 1,929,828,973 | 1,728,052,851 | 332,265,030 | 11,629 | 104,055 | 117,813 |
| 20 | 1,817,182,200 | 1,836,534,794 | 2,061,745,609 | 239 | 102,924 | 118,149 |
| 22.5 | 355,450,930 | 445,159,615 | 1,882,870,790 | 395 | 1,811 | 3,618 |
| 25 | 655,222,874 | 416,072,883 | 349,466,750 | 926 | 2,723 | 5,853 |
| 27.5 | 2,194,229,049 | 1,367,421,458 | 492,207,254 | 868 | 2,468 | 12,072 |
| 30 | 2,587,440,366 | 2,672,284,831 | 1,355,931,485 | 820 | 4,627 | 11,603 |
| 32.5 | 1,817,938,005 | 2,518,792,696 | 3,046,242,779 | 715 | 4,034 | 7,021 |
| 35 | 986,847,330 | 1,205,855,488 | 2,530,303,113 | 306 | 471 | 5,427 |
| 37.5 | 0 | 238,020,337 | 601,433,505 | 149 | 0 | 2,277 |

